# Prevalence of long-term symptoms varies by using different post-COVID-19 definitions in positively and negatively tested adults: the PRIME post-COVID study

**DOI:** 10.1101/2023.07.27.23293244

**Authors:** Demi ME Pagen, Céline JA van Bilsen, Stephanie Brinkhues, Maarten Van Herck, Kevin Konings, Casper DJ den Heijer, Henriëtte LG ter Waarbeek, Martijn A Spruit, Christian JPA Hoebe, Nicole HTM Dukers-Muijrers

## Abstract

**Background:** Long-term symptoms after a SARS-CoV-2 infection (i.e., post-COVID-19 condition or long COVID), constitute a substantial public health problem. Yet, the prevalence remains currently unclear as different case definitions are used, and negatively tested controls are lacking. We aimed to estimate post-COVID-19 condition prevalence using six definitions.

**Methods:** The Prevalence, Risk factors, and Impact Evaluation (PRIME) post-COVID-19 condition study is a population-based sample of COVID-19 tested adults. End 2021, 61,655 adults were invited to complete an online questionnaire, including 44 symptoms plus a severity score (0-10) per symptom. The prevalence was calculated in both positively and negatively tested adults, stratified by time since their COVID-19 test (3-5, 6-11 or ≥12 months ago).

**Results:** In positives (n=7,405; 75.6%), the prevalence of long-term symptoms was between 26.9% and 64.1% using the six definitions, while in negatives (n=2,392; 24.4%) the prevalence varied between 11.4% and 32.5%. The prevalence of long-term symptoms potentially accountable to COVID-19 ranged from 17.9% to 26.3%.

**Conclusion:** There is a (substantial) variation in prevalence estimates by using different definitions as is current practice, showing limited overlap between definitions, indicating that the essential post-COVID-19 condition criteria are still unclear. Including negatives is important to determine long-term symptoms accountable to COVID-19.

Trial registration ClinicalTrials.gov Identifier: NCT05128695.

## Background

Globally, the number of individuals diagnosed with COVID-19 has risen to 660 million by January 2023 (1). Part of all infected individuals report long-term symptoms, that may impede physical and mental limitations, and lead to a loss in work productivity (2–4). These long-term consequences embody a new and growing public health problem. However, the prevalence of long-term symptoms related to COVID-19 (i.e., post-COVID-19 condition or long COVID) to date is unclear, partly due to the diverse terminology used to describe the condition and the lack of an uniform case definition (5).

To improve uniformity, the World Health Organization (WHO) proposed a definition (6), several other health institutes, such as, the UK National Institute for Health and Care Excellence (NICE) (7) and the US Centers for Disease Control (CDC) (8) have proposed other definitions. Definitions used in clinical daily practice are often simplified towards criteria which are more feasible to assess in practice, such as presence of one or more symptoms since infection (9). Recent studies attempted to restrict the number of included symptoms, for example only symptoms that were more present in positively than negatively tested people (10), or that were more severe (11). Also, various durations since infection ranging from several weeks to months were used (8, 12, 13). Until now, it is unknown how the various definitions that are currently in practice relate to each other and to (more complex) post-COVID-19 condition definitions, such as proposed by the WHO.

To construct an adequate case definition, it is crucial to identify direct long-term consequences. It has proven challenging to successfully distinguish direct from possible indirect consequences provoked by the COVID-19 pandemic and preventive measures (14–16). Furthermore, in scientific studies, data on symptoms prior to infection or in negatives controls (as an estimate of background occurrence) is often lacking (17). A Dutch study including test-negative and population controls revealed a substantial proportion of symptom experience (29.8% and 26.0% respectively) in controls, compared to SARS-CoV-2 positive cases (48.5%) (18).

This observational cohort study, called the Prevalence, Risk factors and, Impact Evaluation post-COVID study (PRIME post-COVID), aimed to reveal prevalence and variations in different case definitions. Results provide insight in the complexity assessing post-COVID-19 condition, and providing a reference for future research on long-term symptoms in COVID-19 infected individuals.

## Method

### Study design

In November 2021, the PRIME post-COVID study, with both retrospective and prospective aspects, was initiated. The study design has been published before (19). In brief, a population-based sample of positively and negatively Polymerase Chain Reaction (PCR) tested adults was retrieved from the Dutch public health COVID-19 test registry. All adults with a positive test and a random selected sample of negatively tested adults were selected.

### Participants

In total, 61,655 adults were invited by email to participate. Recorded adults (18 years and older) were invited when they had a valid test result (41,780 positives and 19,875 negatives) and e-mail address. To be classified as negative, no registered positive PCR test was allowed up to the time of participation. Negatively tested invitees were matched (one for each two positives) on age, sex, municipality of residence, and year-quarter of the PCR test. Digital informed consent was obtained before data collection. Consent was also asked to link data on age, sex, and test result from the questionnaire to the public health registry data, for certainty assessment (19).

### Data collection

Data were collected between November 2021 and January 2022 by self-administered questionnaires, by the online MWM2 application of market research platform Crowdtech (ISO 27001 certified). The questionnaire contained demographics (age, sex, comorbidities [per comorbidity whether it was diagnosed before or after testing]), PCR test (date, result), COVID-19 vaccination (number, date(s), type(s) of received vaccines), and experienced symptoms (*“What complaints do you currently have?”*) with severity scores ranging from 0 (no severe symptoms) to 10 (worst symptoms imaginable). There were 44 pre-listed symptoms, based on existing literature (20, 21). All positives were asked whether they felt recovered after infection.

### Outcome variables

The primary outcomes were six definitions of having long-term symptoms after PCR testing (Figure 1). Selection of the definitions was based on WHO recommendation, use in previous studies, and feasibility to be constructed by the study data. A participant was considered a case when reporting:

- Definition 1: ≥1 of all 44 pre-listed symptoms (9);

- Definition 1a: >1 of all 44 pre-listed symptoms;
- Definition 2: ≥1 symptoms that were significantly more often reported in positives than in negatives (in current data) (10);
- Definition 3: ≥1 of the selected symptoms in definition 2 AND with a severity score of ≥5 points (cut-off of 5 was used according to the mean of scores) (11);
- Definition 4: reflects the current WHO case definition: *“a condition that occurs in individuals with a history of probable or confirmed SARS-CoV-2 infection, usually three months from the onset of COVID-19 with symptoms, that last for at least two months, and cannot be explained by an alternative diagnosis’’* (6). A participants was considered a case when reporting ≥1 of the 44 pre-listed symptoms AND the symptoms were present for ≥1 months AND the symptoms were not present before infection AND when no new co-morbidities were reported after the test;
- Definition 5: currently feeling unrecovered (22–25);
- Definition 6: ≥1 of the 44 pre-listed symptoms at 3 months (26), thus reflected for each participant the same time period after positive test (i.e., 3 months).

**Figure 1.**
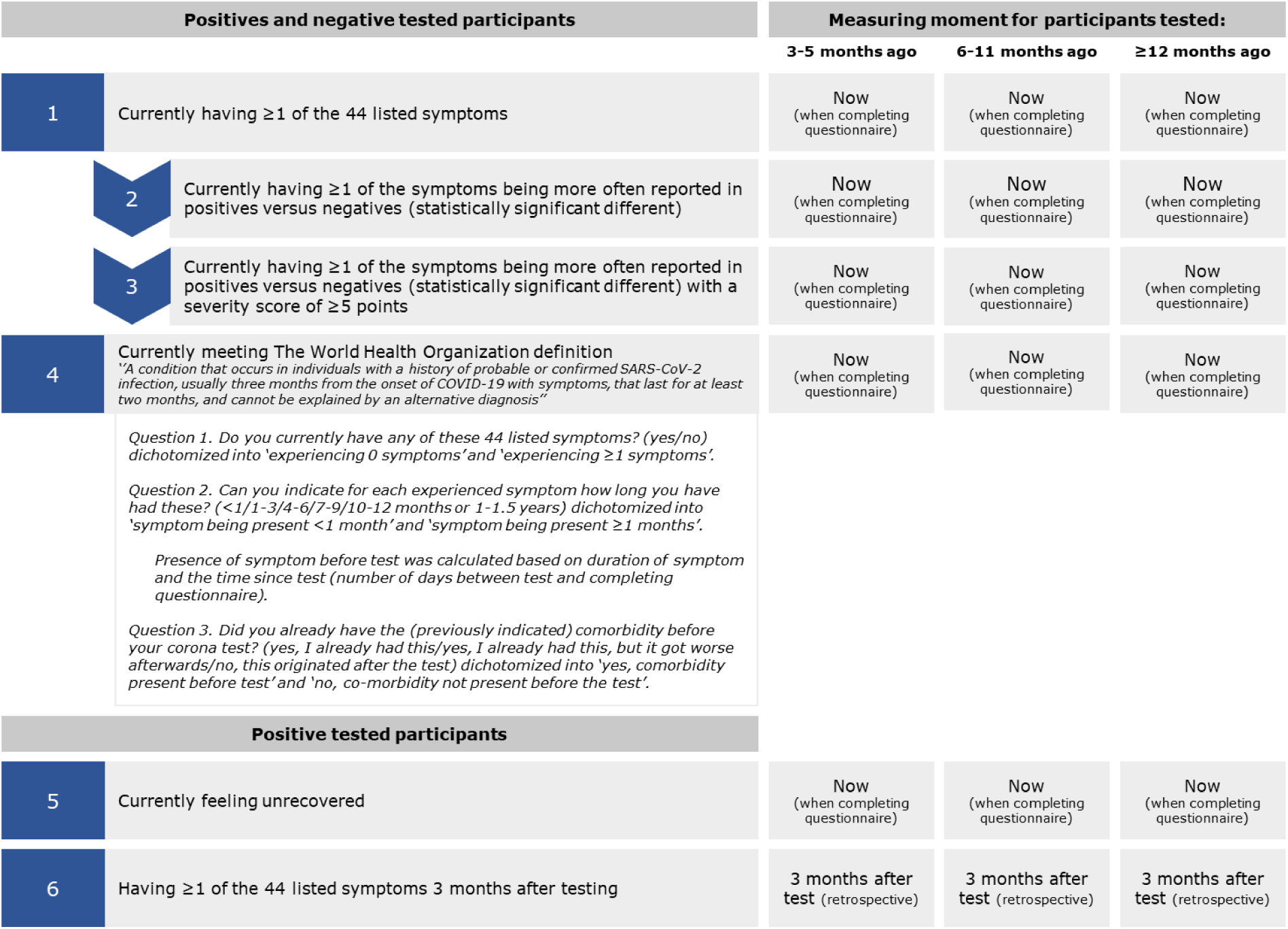
Six definitions of long-term symptoms based on currently experienced symptoms, severity of symptoms, the clinical case definition of post-COVID-19 condition (World Health Organization), currently feeling unrecovered, and experiencing symptoms 3 months after testing, with measuring moments for participants tested 3-5, 6-11 and ≥12 months ago, used in the PRIME post-COVID study

All definitions were constructed for positives, and definition 1-4 also for negatives (as ‘background’ occurrence).

### Time since PCR test

The study population was divided into categories based on time since PCR test (3-5∼Delta variant, 6-11∼Alpha variant, and ≥12 months∼Wuhan variant). Respondents who had a duration shorter than 3 months were excluded (i.e., not indicative for long-term symptoms).

### Statistical analysis

To improve the likelihood for representativeness of the study population relative to the invitees, data were weighted by age categories, sex, and year-quarter of PCR test (positives and negatives separately). Unweighted numbers and percentages (u%) and weighted percentages (w%) were presented. Chi-square tests were used to compare symptoms between all positives and negatives and positives and negatives who met definitions 1 and 4 (definitions 2 and 3 are not representative of all 44 pre-listed symptoms and definitions 5 and 6 were unavailable for negatives). The proportion of participants with the outcomes (for each definition separately) was estimated and 95% Confidence Intervals were calculated, stratified by time since the test: 3-5, 6-11 and ≥12 months ago. The range of prevalence estimates was presented for the three test window groups. Logistic regression analyses were performed to test whether the presence of long-term symptoms differed between the test window groups. The proportion of long-term symptoms potentially accountable to a positive SARS-CoV-2 test (as estimate for direct rather than indirect impact) was calculated by subtracting the observed prevalence in negatives from the observed prevalence in positives. We stress ‘potentially accountable’ since we acknowledge that we cannot rule out that (part of) negatively tested participants might have been infected, but not included in the registry. In sensitivity analyses, negatively tested participants reporting SARS-CoV-2 antibodies (type not specified) prior to vaccination were excluded. To compare inter-relations between the definitions used in practice (definition 1, 5, and 6 were selected for clarity and readability) and with the WHO case definition, three Venn diagrams were constructed for each of the three test window groups. Statistical analyses were performed using Statistical Package for Social Sciences (SPSS; version 27.0, IBM, Armonk, USA). A p-value <0.001 was considered statistically significant applying Bonferroni correction due to multiple testing (27).

### Ethical statement

The Medical Ethical Committee of Maastricht University Medical Centre+ waived this study (METC2021-2884), as the Medical Research Involving Human Subjects Act (WMO) did not apply. This study was registered at Clinical Trials.gov Protocol Registration and Results System (NCT05128695).

## Results

Of the 61,655 invitees, 12,453 (20.2%) participants provided the minimal data of whom 9,797 (78.7%) were PCR tested ≥3 months ago and thereby eligible for inclusion (Figure 2).

**Figure 2.**
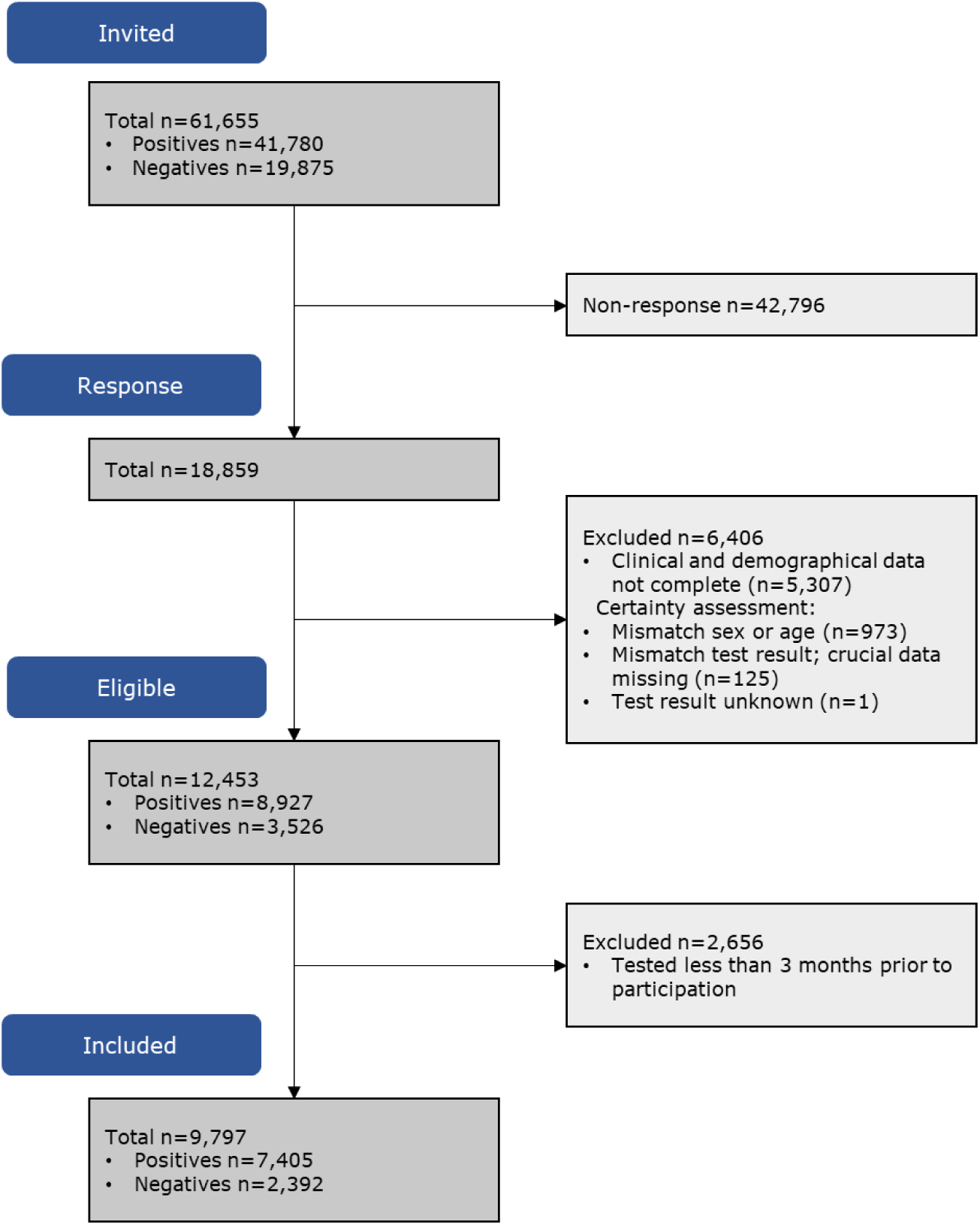
Flowchart of invitees, respondents, exclusion criteria, participants eligible for inclusion, and study population included in the analysis of the PRIME post-COVID study

Of the included participants, 7,405 (75.6%) had tested positive, 2,392 (24.4%) negative, and 1,367 (14.0%) were tested 3-5 months ago, 6,402 (65.3%) 6-11 months ago, and 2,028 (20.7%) ≥12 months ago (Table 1). The share of positives was 61.9%, 80.0%, and 71.0% for the test window groups, respectively. For participants tested ≥6 months ago, negatives were more often men compared to positives (p<0.001) (Table 1). Overall, negatives were older than positives (p<0.001).

**Table 1.**
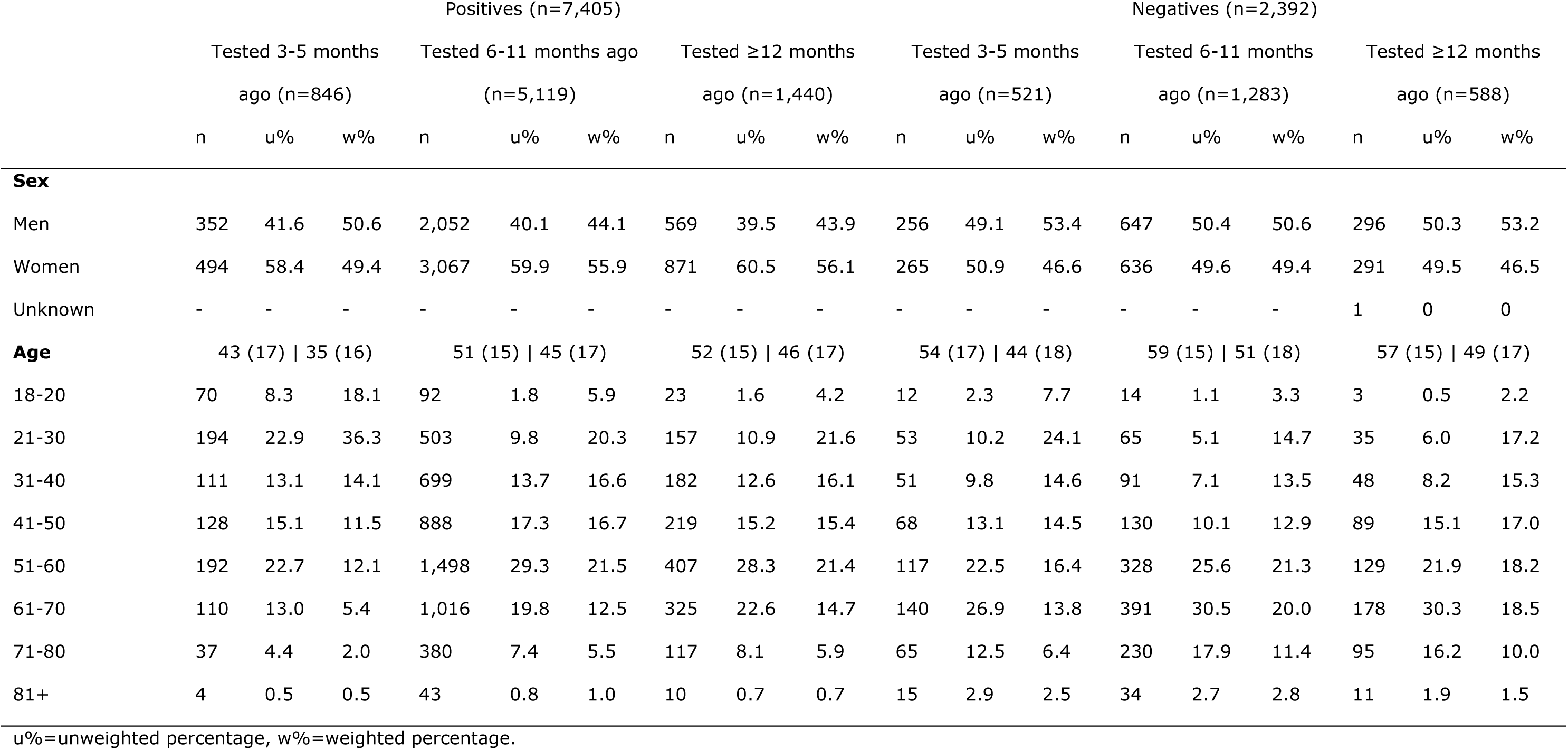
Participant characteristics for the positively and negatively tested adults stratified for time since test.

The majority of all participants had received at least one vaccine dose (80.6%, 91.3%, and 91.7% of positives and 98.0%, 96.9%, and 97.1% of negatives, for the test window groups, respectively).

The proportion of negatives who experienced ≥1 symptoms around the moment of their test was lower (p<0.001) compared to positives. In total, 19.6% (n=1,450) of positives reported >10 symptoms and 1.3% (n=99) were hospitalized during acute infection. Comorbidities reportedly present prior to the PCR test was overall comparable between positives and negatives. Current comorbidities were reported more often in negatives than positives, tested 3-5 months ago (44.3% negatives, 32.5% positives; p<0.001).

### Description of symptoms significantly more often reported in positives than negatives

Of the 44 pre-listed symptoms, 24 were more often reported in positives than in negatives, including amnesia, brain fog, chest tightness, concentration difficulties, confusion, cough, dizziness, fatigue, hair loss, headache, heart palpitations, increased resting heart rate, irritability, joint pain, loss/change of smell, loss/change of taste, mucus, muscle pain and weakness, pain between shoulder blades, pain or burning sensation in the lungs, shortness of breath, sleeping problems, tinnitus, and voice difficulties. These symptoms were used in definitions 2 and 3 (see methods).

### Reported symptoms in positives and negatives who met the case definitions

Negatives who met the case definitions 1 or 4 significantly more often experienced general cold symptoms (i.e., earache, sneezing, runny nose, cold), vomiting, and dreariness/depression, compared to positives who met definitions 1 or 4 (Table 2). Symptoms significantly more often reported in positives than negatives meeting definitions 1 or 4 were chest pressure, hair loss, amnesia, loss/change of taste, loss/change of smell, shortness of breath, concentration difficulties, and fatigue (Table 2). The proportion of experienced loss/change of smell (20.2% for definition 1 and 24.1% for definition 4) and fatigue (16.8% for definition 1 and 8.2% for definition 4) described the largest difference between positives and negatives (Table 2).

**Table 2.** Proportion of symptoms experienced by positives and negatives meeting four long-term symptom definitions.

| Experienced symptoms (w%) | Definition 1.<br>(≥1 of all symptoms) n=4,488* |  |  | Definition 4. (WHO)<br>n=3,095* |  |  | Definition 5.<br>(unrecovered)<br>n=2,428* | Definition 6.<br>(≥1 of all symptoms<br>at 3 months)<br>n=4,438* |
| --- | --- | --- | --- | --- | --- | --- | --- | --- |
|  | Negatives | Positives | Diff. | Negatives | Positives | Diff. | Positives | Positives |
|  | (n=721) | (n=3,767) |  | (n=345) | (n=2,322) |  |  |  |
| Loss/change of smell | 3.1 | 23.3 | 20.2† | 3.5 | 27.6 | 24.1† | 28.3 | 19.2 |
| Fatigue | 31.9 | 48.7 | 16.8† | 45.2 | 53.4 | 8.2 | 57.8 | 38.7 |
| Shortness of breath | 8.1 | 20.7 | 12.6† | 9.0 | 22.1 | 13.1† | 27.9 | 16.6 |
| Concentration difficulties | 10.6 | 23.0 | 12.4† | 15.4 | 24.8 | 9.4† | 30.4 | 19.0 |
| Loss/change of taste | 3.3 | 15.7 | 12.4† | 3.5 | 18.3 | 14.8† | 20.4 | 12.9 |
| Amnesia | 4.7 | 11.7 | 7.0† | 6.1 | 12.3 | 6.2† | 15.4 | 9.5 |
| Chest pressure | 2.4 | 6.9 | 4.5† | 2.6 | 6.9 | 4.3 | 9.3 | 5.5 |
| Hair loss | 3.6 | 7.4 | 3.8† | 6.1 | 8.6 | 2.5 | 8.3 | 5.9 |
| Headache | 13.2 | 16.4 | 3.2 | 18.9 | 16.3 | -2.6 | 19.0 | 12.6 |
| Brain fog | 5.0 | 8.2 | 3.2 | 6.1 | 8.4 | 2.3 | 10.9 | 6.7 |
| Pain between shoulder blades | 4.7 | 7.0 | 2.3 | 5.8 | 7.3 | 1.5 | 8.8 | 5.5 |
| Palpitations | 4.3 | 6.4 | 2.1 | 6.1 | 6.4 | 0.3 | 8.4 | 5.2 |
| Increased resting heart rate | 2.6 | 4.7 | 2.1 | 3.8 | 4.8 | 1.0 | 6.6 | 3.7 |
| Confusion | 1.2 | 2.9 | 1.7 | 1.2 | 2.9 | 1.7 | 4.1 | 2.3 |
| Pain or burning sensation in the lungs | 1.2 | 2.8 | 1.6 | 1.2 | 3.0 | 1.8 | 4.0 | 2.2 |
| Irritability | 8.3 | 9.9 | 1.6 | 13.0 | 9.9 | -3.1 | 13.1 | 8.1 |
| Muscle pain or weakness | 9.7 | 11.0 | 1.3 | 13.7 | 11.2 | -2.5 | 14.0 | 8.6 |
| Voice difficulties | 3.6 | 4.5 | 0.9 | 5.2 | 4.6 | -0.6 | 5.7 | 3.3 |
| Diarrhoea | 1.9 | 2.7 | 0.8 | 2.9 | 2.8 | -0.1 | 3.4 | 2.1 |
| Elevated body temperature | 0.7 | 1.1 | 0.4 | 0.6 | 0.9 | 0.3 | 1.5 | 0.8 |
| Heat flushes | 1.4 | 1.8 | 0.4 | 2.6 | 1.9 | -0.7 | 2.0 | 1.5 |
| Fever | 0.1 | 0.2 | 0.1 | 0.0 | 0.0 | 0.0 | 0.2 | 0.1 |
| Sudden weight loss | 0.8 | 0.9 | 0.1 | 1.7 | 0.9 | -0.8 | 1.2 | 0.8 |
| Burning sensation in the trachea | 2.1 | 2.1 | 0.0 | 2.3 | 2.4 | 0.1 | 3.0 | 1.7 |
| Stomach ache | 2.9 | 2.6 | -0.3 | 3.5 | 2.8 | -0.7 | 3.0 | 2.1 |
| Dizziness | 7.1 | 6.8 | -0.3 | 10.2 | 6.9 | -3.3 | 9.1 | 5.5 |
| Sleeping problems | 16.6 | 16.3 | -0.3 | 20.3 | 16.8 | -3.5 | 20.3 | 13.1 |
| Loss of appetite | 2.6 | 2.2 | -0.4 | 4.1 | 2.3 | -1.8 | 3.1 | 1.8 |
| Tinnitus | 8.5 | 8.1 | -0.4 | 7.8 | 7.8 | 0.0 | 8.5 | 6.3 |
| Skin rash/red spots on toes or feet | 1.5 | 1.0 | -0.5 | 2.9 | 0.9 | -2.0† | 1.1 | 0.8 |
| Nerve pain | 3.3 | 2.8 | -0.5 | 4.4 | 2.9 | -1.5 | 3.6 | 2.2 |
| Vomiting | 0.8 | 0.2 | -0.6† | 1.4 | 0.1 | -1.3† | 0.2 | 0.1 |
| Fear | 5.1 | 4.4 | -0.7 | 6.7 | 3.7 | -3.0 | 5.7 | 3.5 |
| Eye difficulties | 5.8 | 5.0 | -0.8 | 7.2 | 5.5 | -1.7 | 6.4 | 4.0 |
| Nausea | 3.5 | 2.7 | -0.8 | 4.4 | 2.3 | -2.1 | 3.2 | 2.1 |
| Joint pain | 11.4 | 10.4 | -1.0 | 12.5 | 10.1 | -2.4 | 12.0 | 8.2 |
| Coughing up mucus | 11.1 | 10.0 | -1.1 | 12.2 | 9.6 | -2.6 | 10.4 | 6.5 |
| Sore throat | 9.0 | 7.6 | -1.4 | 9.3 | 6.4 | -2.9 | 7.2 | 4.9 |
| Earache | 3.3 | 1.8 | -1.5 | 4.6 | 1.6 | -3.0† | 2.0 | 1.4 |
| Other | 5.3 | 3.6 | -1.7 | 5.8 | 3.3 | -2.5 | 3.8 | 2.9 |
| Coughing | 17.4 | 14.0 | -3.4 | 17.7 | 12.1 | -5.6 | 13.1 | 8.9 |
| Dreariness/depression | 12.5 | 9.1 | -3.4 | 15.7 | 8.5 | -7.2† | 10.2 | 7.2 |
| Sneezing | 15.8 | 7.7 | -8.1† | 15.7 | 6.5 | -9.2† | 6.3 | 4.8 |
| Cold | 30.0 | 19.7 | -10.3† | 26.1 | 15.6 | -10.5† | 14.0 | 11.9 |
| Runny nose | 23.3 | 11.9 | -11.4† | 23.5 | 9.7 | -13.8† | 8.3 | 7.6 |

\* Number of participants meeting the definition.
† Significantly different between positives and negatives meeting the definitions ( $p \leq 0.001$ ).
w%=weighted percentage, diff=difference calculated by subtracting the proportion of the experienced symptom in the negatives from the proportion of the experienced symptom in the positives. Negative values represent symptoms more often present in negatives; positive values represent symptoms more frequently reported in positives.
Definition 1. Currently reporting $\geq 1$ of the 44 pre-listed symptoms; Definition 4. World Health Organization post-COVID-19 condition case-definition; Definition 5. Currently feeling unrecovered since PCR test; and Definition 6. Reporting $\geq 1$ of the 44 pre-listed symptoms 3 months after testing.

The most reported symptoms in positives who met definition 5 and 6 were fatigue, loss/change of smell, shortness of breath, concentration difficulties, change/loss of taste, and sleeping problems (Table 2).

### Prevalence of six long-term symptom definitions in positives and negatives

In positives, the prevalence of long-term symptoms ranged between 26.9% to 64.1% for all definitions (Figure 3): the prevalence ranged (by test window group) between 47.6%-53.1% for definition 1 (≥1 of all symptoms), 34.9%-39.2% by definition 1a (Supplementary Table 1), 41.1%-47.0% for definition 2 (different symptoms positives-negatives), 33.4%-37.9% for definition 3 (different symptoms positives-negatives plus severity), 34.7%-39.0% for definition 4 (WHO), 26.9%-34.4% for definition 5 (unrecovered), and 47.4%-64.1% for definition 6 (≥1 of all symptoms at 3 months). In negatives, the prevalence was between 29.0%-32.5%, 18.5%-22.6%, 14.0%-18.8%, and 11.4%-19.3%, for definitions 1 – 4, respectively (Figure 3). Excluding negatives with SARS-CoV-2 antibodies prior to vaccination (n=95) showed comparable results (Supplementary Table 2).

**Figure 3.**
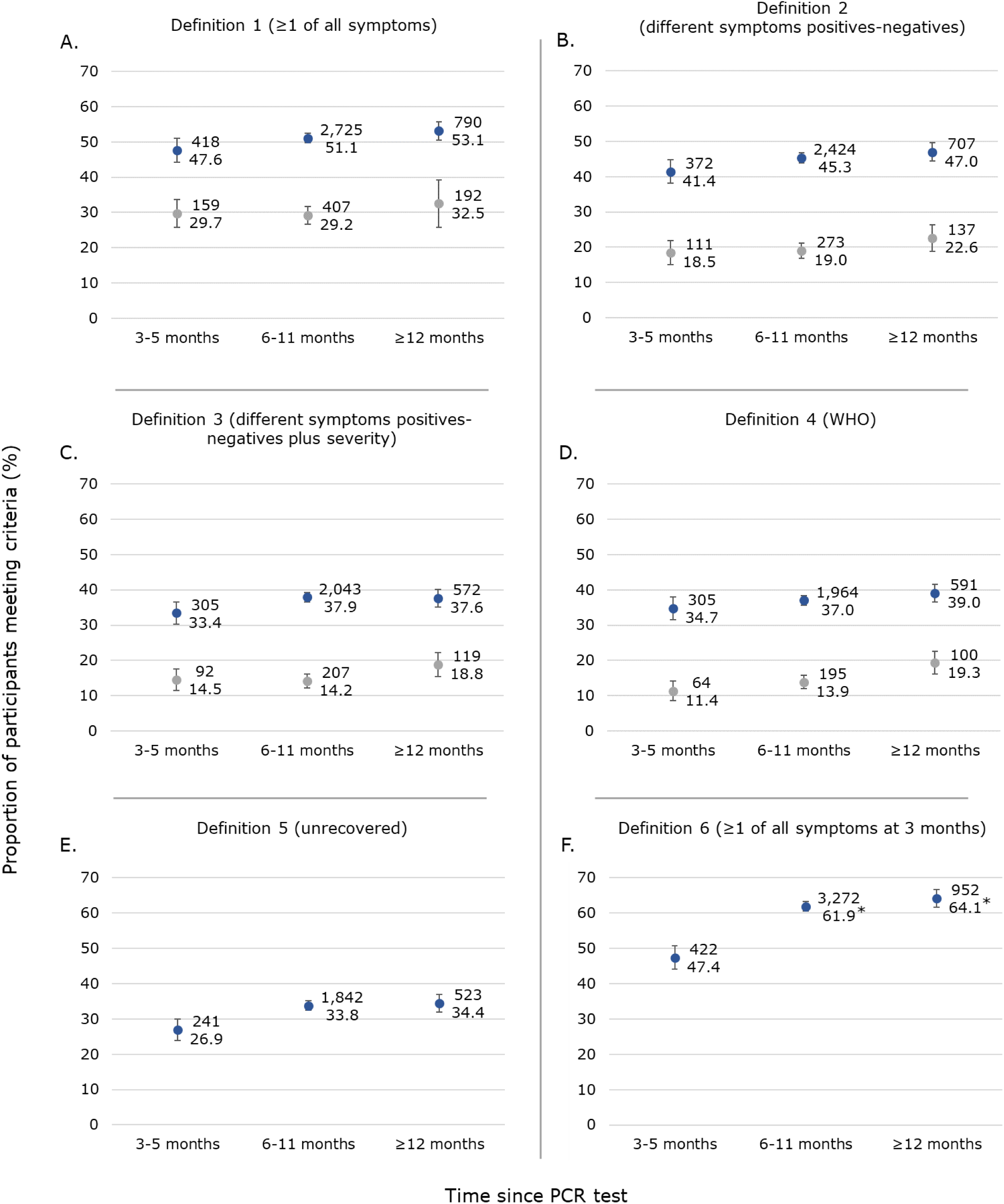
Weighted prevalence estimates and 95% confidence intervals for six long-term symptom definitions in positives and negatives stratified for time since Polymerase Chain Reaction (PCR) test. 3A. Definition 1. Currently reporting ≥1 of the 44 pre-listed symptoms; 3B. Definition 2. Currently reporting ≥1 of the 24 symptoms more often reported in positives versus negatives; 3C. Definition 3. Reporting ≥1 of the 24 symptoms more often reported in positives versus negatives with a severity score of ≥5; 3D. Definition 4. Meeting the World Health Organization definition; 3E. Definition 5. Currently feeling unrecovered since PCR test; and 3F. Definition 6. Reporting ≥1 of the 44 pre-listed symptoms 3 months after testing. * indicated significant differences in prevalence estimates compared to participants tested 3-5 months ago. Dominant virus variants in overlapping periods were Wuhan strain between March-December 2020; Alpha strain between December 2020-July 2021; Delta strain between July-December 2021.

Prevalence estimates for definitions 1-5 did not differ between test window groups. However, for definition 6 (≥1 of all symptoms 3 months) the prevalence was significantly higher in positives tested 6-11 months (61.9%) and ≥12 months (64.1%) ago, compared to positives tested 3-5 months (47.4%) ago.

### Long-term symptoms potentially accountable to COVID-19

The long-term symptoms proportion difference between positives and negatives – thus long-term symptoms potentially accountable to COVID-19 – ranged between 17.9% and 26.3% for all definitions: between 17.9%-21.9% for definition 1 (≥1 of all symptoms), 22.8%-26.3% for definition 2 (different symptoms positives-negatives), 18.2%-23.7% for definition 3 (different symptoms positives-negatives plus severity), and between 21.4%-23.3% for definition 4 (WHO) (Figure 4).

**Figure 4.**
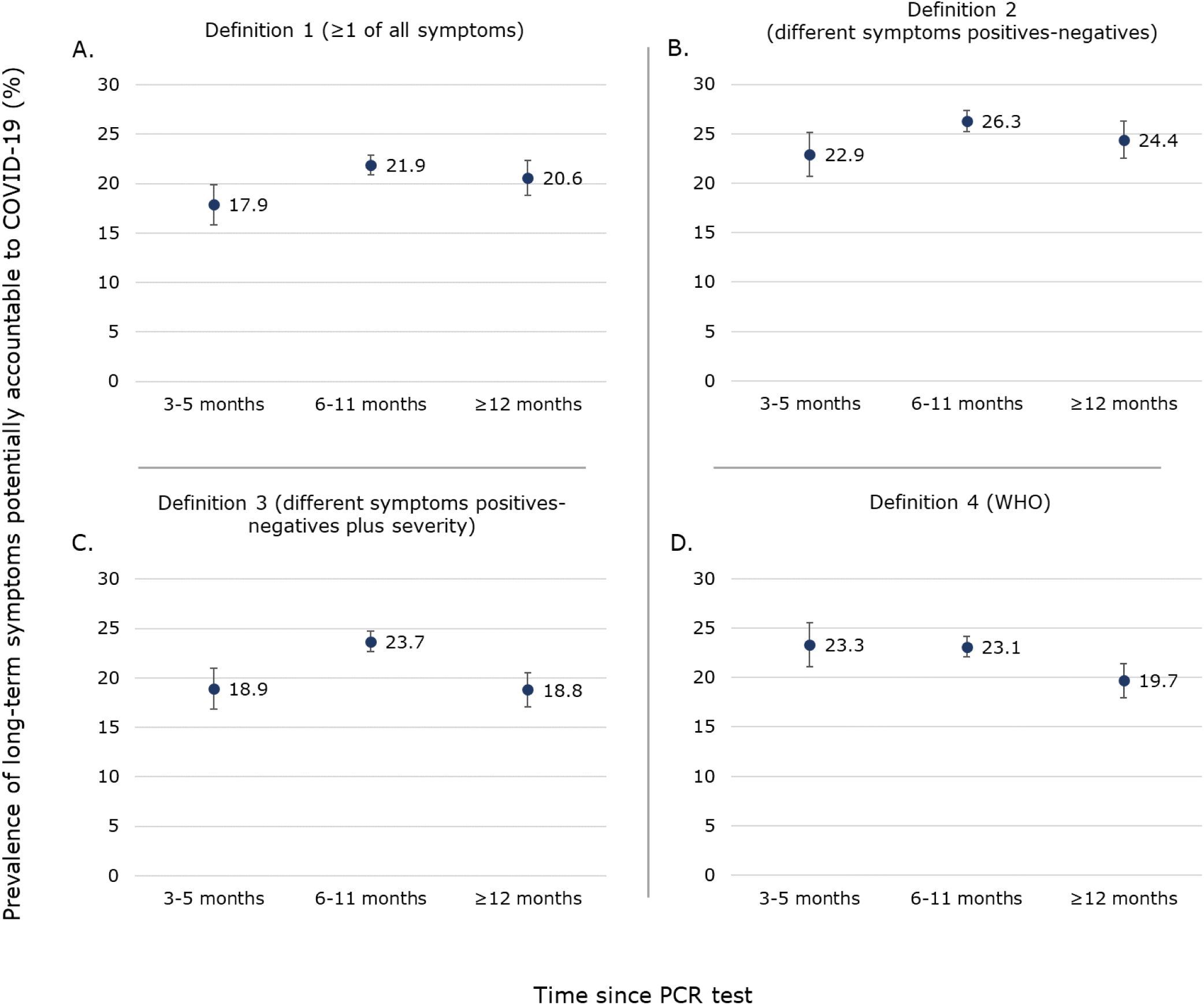
Weighted prevalence estimates and 95% confidence intervals for long-term symptoms potentially accountable to COVID-19 using four definitions, stratified for time since Polymerase Chain Reaction (PCR) test. 4A. Definition 1. Currently reporting ≥1 of the 44 pre-listed symptoms; 4B. Definition 2. Currently reporting ≥1 of the 24 symptoms more often reported in positives versus negatives; 4C. Definition 3. Reporting ≥1 of the 24 symptoms more often reported in positives versus negatives with a severity score of ≥5; 4D. Definition 4. Meeting the World Health Organization definition. Potentially accountable prevalence was calculated by subtracting the observed prevalence in negatives from the observed prevalence in positives.

### Overlap between case definitions in positives

In total, 519 (61.4%), 3,567 (69.7%), and 1,043 (72.4%) positives tested 3-5, 6-11, and ≥12 months ago, respectively, met at least one of the six outcome case definitions.

Of these, the majority of the participants with long-term symptoms, met definition 1 (≥1 of all symptoms; 80.5% tested 3-5 months ago, 76.4% tested 6-11 months ago, and 75.7% tested ≥12 months ago) and definition 6 (≥1 of all symptoms at 3 months; 85.2% tested 3-5 months ago, 91.7% tested 6-11 months ago, and 91.3% tested ≥12 months ago) (Figure 5).

**Figure 5.**
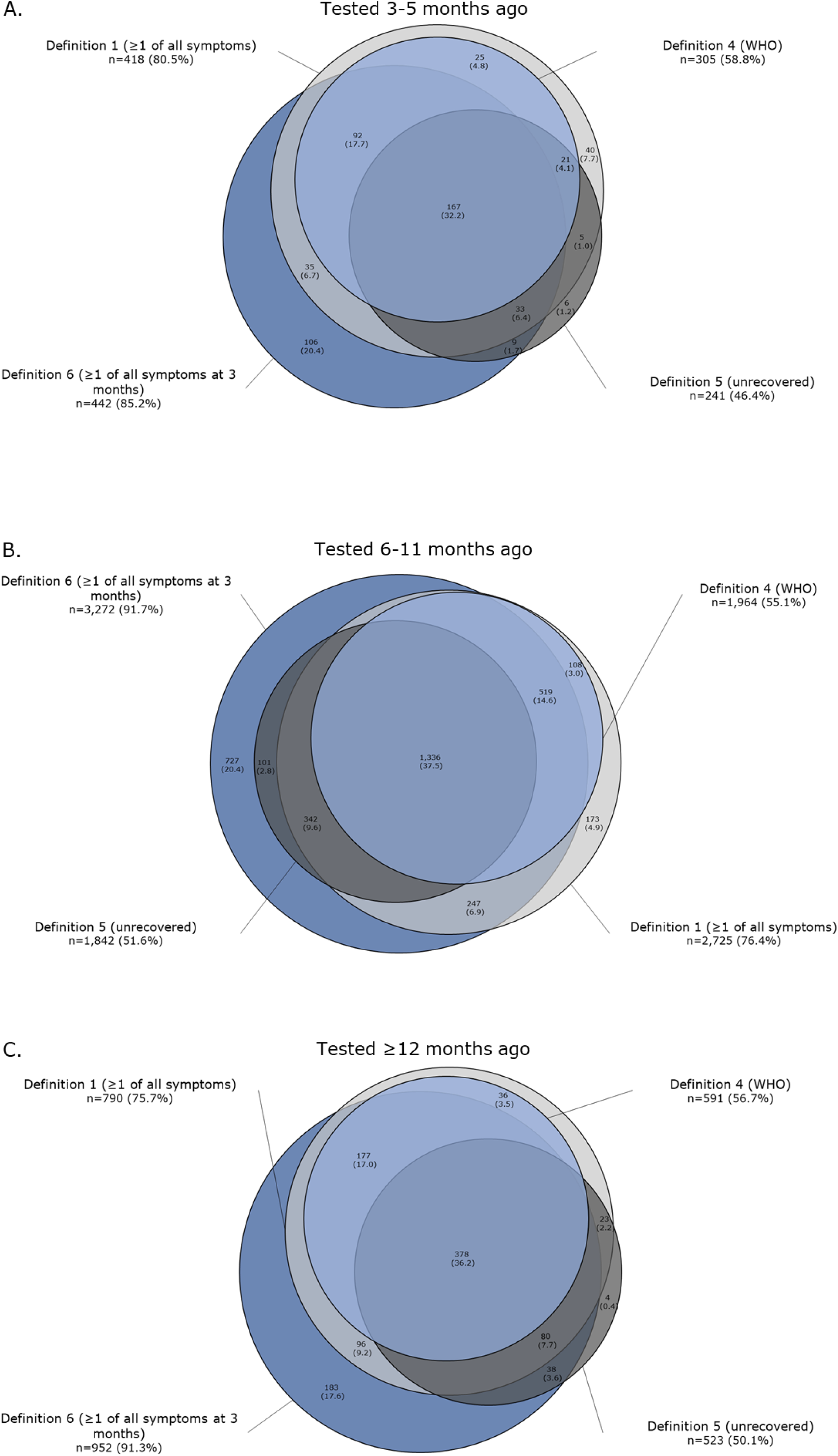
Venn diagrams of four long-term symptom definitions in positively tested participants. Definition 1. Currently Reporting ≥1 of the 44 pre-listed symptoms; Definition 4. Meeting the World Health Organization definition; Definition 5. Currently feeling unrecovered since PCR test; and Definition 6. Reporting ≥1 of the 44 pre-listed symptoms 3 months after testing. Percentages represent the proportion of participants meeting the definitions compared to all positives who met at least one of the definitions (n=519 for participants tested 3-5 months ago, n=3,567 for participants tested 6-11 months ago, and n=1,043 for participants tested ≥12 months ago).

About half of the participants with long-term symptoms met definition 4 (WHO) and 5 (unrecovered) (Figure 5). The overlap between the definitions depicted in the Venn diagrams was similar for the different test window groups.

The proportion of participants who met the WHO definition and would also meet definitions used in clinical practice (definitions 1, 5, and 6), was 100.0%, 67.9%, and 93.4% for definitions 1, 5, and 6, respectively. The proportion of participants not meeting the WHO definition (definition 4), and similarly not meeting definitions 1, 5, and 6, was 76.4%, 85.4% and 56.5% respectively (Table 3).

**Table 3.** Comparison between the definitions used in daily clinical practice (definitions 1, 5, and 6) and the WHO definition.

|  |  | Definition 4 (WHO) |  |
| --- | --- | --- | --- |
|  |  | Met definition | Did not meet definition |
| Tested 3-5 months ago (n=846) |  | (n=305) | (n=541) |
| Definition 1 (≥1 of all symptoms) | Met definition (n=418) | 100.0% |  |
|  | Did not meet definition (n=428) |  | 79.1% |
| Definition 5 (unrecovered) | Met definition (n=241) | 61.6% |  |
|  | Did not meet definition (n=605) |  | 90.2% |
| Definition 6 (≥1 of all symptoms at 3 months) | Met definition (n=422) | 84.9% |  |
|  | Did not meet definition (n=424) |  | 69.9% |
| Tested 6-11 months ago (n=5,119) |  | (n=1,964) | (n=3,155) |
| Definition 1 (≥1 of all symptoms) | Met definition (n=2,725) | 99.9% |  |
|  | Did not meet definition (n=2,394) |  | 75.9% |
| Definition 5 (unrecovered) | Met definition (n=1,842) | 69.4% |  |
|  | Did not meet definition (n=3,277) |  | 84.8% |
| Definition 6 (≥1 of all symptoms at 3 months) | Met definition (n=3,272) | 94.5% |  |
|  | Did not meet definition (n=1,847) |  | 55.1% |
| Tested ≥12 months ago (n=1,440) |  | (n=591) | (n=849) |
| Definition 1 (≥1 of all symptoms) | Met definition (n=790) | 100.0% |  |
|  | Did not meet definition (n=650) |  | 76.6% |
| Definition 5 (unrecovered) | Met definition (n=523) | 66.0% |  |
|  | Did not meet definition (n=917) |  | 84.3% |
| Definition 6 (≥1 of all symptoms at 3 months) | Met definition (n=952) | 93.9% |  |
|  | Did not meet definition (n=488) |  | 53.2% |

WHO=world health organization.
Percentages represent proportion of participants who met definition 4 (WHO) and also met definitions 1 (≥1 of all symptoms), 5 (unrecovered), and 6 (≥1 of all symptoms at 3 months), OR the proportion of participants who did not meet definition 4 (WHO) and also did not meet definitions 1 (≥1 of all symptoms), 5 (unrecovered), and 6 (≥1 of all symptoms at 3 months).

## Discussion

Results of the PRIME post-COVID cohort study demonstrate that loss/change of smell, fatigue, loss/change of taste, shortness of breath, and concentration difficulties reflect the most pronounced long-term symptoms. Prevalence estimates of long-term symptoms after PCR positive testing show a large range from 26.9% to 53.1%, depending on the definition used in science and care. Variation was also recorded by moment of repot, demonstrating 64.1% based on retrospective assessment of symptoms present 3 months after testing. Participants who tested negative also reported symptoms, with substantial proportions ranging from 11.4% to 32.5%. Accounting for symptoms in negatives, the prevalence of long-term symptoms potentially accountable to a SARS-CoV-2 infection ranged from 17.9% to 26.3%.

Including negatives enables an estimation of the proportion of direct consequences of COVID-19 infection. Yet, we acknowledge that we cannot rule out that (part of) tested negatives might have been infected (e.g., due to undiagnosed infection at the time of questionnaire completion), but unknown in our dataset. Despite the varying prevalence estimates when using different definitions, the prevalence potentially accountable to COVID-19 is within reasonable range across the various case definitions (between 17.9% and 26.3%). These findings suggest that the inclusion of a control group of negatively tested participants can be of great value when estimating post-COVID-19 condition prevalence, independent of definition used.

Comparing prevalence estimates of our study with previous literature is challenging, due to unstandardized definitions of relevant symptoms after testing or infection. Prevalence estimates from previous studies ranged from 6.0% to 80.0% in positives, 26.0% to 53.4% in negatives, and the proportion accountable to COVID-19 ranged from 12.7% to 25.2% (10, 11, 25, 28–31). The range in estimated accountable symptoms is comparable to our findings.

The type of symptoms and the proportion of positives and negatives who reported to experience those symptoms can be very different when using different case definitions. For example, the difference between experiencing fatigue in positively versus negatively tested participants meeting definition 1 (≥1 of all symptoms) was twice as high compared to the difference in experienced fatigue between positively and negatively tested participants who met definition 4 (WHO). This clearly illustrates the difficulty to define a core set of symptoms, needed to reach the desired consensus.

Results of our study suggest that defining the presence of long-term symptoms based on any symptoms, likely overestimates prevalence of post-COVID-19 condition. Selecting only symptoms significantly more often reported in positives than negatives and including a degree of severity results in lower prevalence estimates. While this might result in more realistic estimates, it should be noted that such definition also might still be an underestimation, due to potentially relevant unmeasured symptoms. Recent studies included symptom severity in the case definition (10, 11), suggesting this might have an added value when studying post-COVID-19 condition. Our results provide a range of prevalence estimates based on different definitions, highlighting the complexity of this condition and measurements, and providing a reference for future research. Focussing on a single definition will cause cases to be missed, exemplified in our Venn diagram and comparison with the WHO definition. Only including the WHO definition when studying post-COVID-19 condition will identify about half (55.1% to 58.8% for different test window groups) of all likely cases (based on definitions 1-3, 5, and 6). Thereby, we were able to compare more simple (in practice) definitions with each other and the current WHO definition. Yet, this also indicates that the search for an adequate case definition is not completed, and finding the essential criteria for defining post-COVID-19 condition is still a challenge.

No differences in prevalence estimates were observed test window groups. Only for definition 6 (constructed by retrospective reporting; ≥1 of all symptoms at 3 months) prevalence was significantly higher in participants tested 6-11 and ≥12 months ago, compared to participants tested 3-5 months ago. Whether this is due to potential (recall)bias, virus variant (overlapping with the time periods since test used in the current study) or vaccination status has to be further explored. Currently, only data of the baseline recruitment (cross-sectional) is available, eliminating the opportunity to study prevalence estimates in more recent calendar times and individual trajectories over time. Future analysis using longitudinal data collected within the PRIME post-COVID study will provide more insight into this issue.

### Strengths and limitations

The major strength of this study is the population-based sample, also including positives who experienced a mild infection. Additionally, we were able to include a considerable portion of negatives, creating the opportunity to study long-term symptoms experienced in the general population and the proportion of long-term symptoms potentially accountable to COVID-19. Nevertheless, negatives might have been infected, but not tested, due to limited test possibilities in the beginning of the pandemic, lack of indication for testing (i.e., asymptomatic nature of the infection) or lack of intention to test. This might have affected our comparison of symptoms and prevalence estimates between positives and negatives, possibly leading to some over-correction. Nevertheless, sensitivity analyses excluding negatives with SARS-CoV-2 antibodies prior to vaccination showed comparable results. Regarding the validity, we were able to check questionnaire data with public health registry data, resulting in more confirmed certainty of our data (i.e., age, sex, test result and test date).

First, possible limitations in generalizability have to be discussed. Since all positively tested adults in the registry were invited, the invitees were representative of all positives recorded in the Dutch public health test registry (in our study region). Nevertheless, generalizability of the individuals in the registry regarding all positive adults in the population might not be optimal. For example, the number of individuals suffering severe illness was limited, possibly resulting in an underestimation of post-COVID-19 condition in our sample (32). Besides, a limited number of asymptomatic positives were included, due to lack of motivation for testing and thus no registration in the public health registry. Still, a part of positively and negatively tested participants did not experience any symptoms when tested, meaning that testing – and registration – was probably indicated by other factors (e.g., source and contact tracing). It is expected that the lack of asymptomatic positives has a limited effect, as post-COVID-19 condition prevalence is estimated to be very limited in this population.

Negative invitees were comparable to positive invitees in key characteristics, however not necessarily to all negatives in the registry. Invited negatives included less women (50.1% versus 55.6%), less adults tested in the second quarter of 2020 (18.3% versus 23.4%), and more adults aged 51-70 years (34.1% versus 24.1%), compared to all negatives in the registry. We acknowledge the limited generalizability of negative participants to negatives in the registry, but recognized the value of comparable characteristics between positive and negative invitees.

Second, there is a possibility of selection bias overall, as not all invitees participated. Elderly who are less digitally skilled were probably more likely to decline participation, resulting in an underrepresentation of these participants. However, we tried to limit the influence of selection bias on our results by weighing participants back to invitees on key characteristics (age, sex, year-quarter of testing, test result). Despite, we observed negatives to be more often men (when tested ≥6 months ago) and older of age, which we will take into account in future analyses.

In conclusion, prevalence estimates of long-term symptoms after infection vary widely, between 26.9% to 53.1% when using different definitions based on current symptoms (i.e., measuring moment now) and 64.1% when using a definition based on retrospective assessment of symptoms (i.e., 3 months after PCR test) in positives. This highlights the importance of formulating an adequate post-COVID condition definition to reliably estimate the prevalence of long-term symptoms accountable to a positive SARS-CoV-2 test. Taking severity of experienced symptoms into account facilitates specification of prevalence estimates by only including symptoms having at least moderate impact. Negatives or population controls are important to determine long-term symptoms accountable to COVID-19, preventing overestimation of the prevalence. Furthermore, there is limited overlap between different long-term symptom definitions, indicating that the essential criteria for defining post-COVID-19 condition are still unclear. Future studies should focus on risk factors predisposing certain individuals, to help focus on relevant subgroups in practice, for the clinic or the general population.

## Data Availability

Data cannot be shared publicly because the data contains potentially identifying patient information. Data are available on request from the head of the data-archiving South Limburg Public Health Service (contact via) for researchers who meet the criteria for access to confidential data.

## Author contributions

DP, CvB, SB, MVH, KK, CdH, HtW, MS, CH, ND designed the study. DP, CvB and SB actively participated in data collection. DP performed the data analysis and wrote the first draft of the manuscript. CvB, MVH, KK, CdH, and ND supervised data analysis and collection. All authors were involved in data interpretation, revised the manuscript critically for important intellectual content, approved the final version, and agreed to be accountable for all aspects of the manuscript.

## Acknowledgements

We gratefully acknowledge LCJ Steijvers, CPB Moonen, S Mujakovic, AW Vaes and N Bouwmeester-Vincken for their valuable contribution to the development of the questionnaire, and CPB Moonen for her participation in data collection.

