## Supplementary Table 1 for "Prevalence of long-term symptoms varies by using different post-COVID-19 definitions in positively and negatively tested adults: the PRIME post-COVID study"

Supplementary Table 1. Long-term symptom definition 1a (having >1 of the 44 pre-listed symptoms) for positively tested adults

|  | Positives (n=7,405) | | | | | | | | |
| --- | --- | --- | --- | --- | --- | --- | --- | --- | --- |
|  | Tested 3-5 months ago (n=846) | | | Tested 6-11 months ago (n=5,119) | | | Tested ≥12 months ago (n=1,440) | | |
|  | n | u% | w% | n | u% | w% | n | u% | w% |
| Definition 1a. (>1 of all symptoms) | 312 | 36.9 | 34.9 | 2,082 | 40.8 | 38.4 | 596 | 41.6 | 39.2 |
| u%=unweighted percentage, w%=weighted percentage. | | | | | | | | | |
