## Supplementary Table 2 for "Prevalence of long-term symptoms varies by using different post-COVID-19 definitions in positively and negatively tested adults: the PRIME post-COVID study"

Supplementary Table 2. Long-term symptom definitions for negatively tested adults when excluding negatives reporting SARS-CoV-2 antibodies prior to vaccination

|  | Negatives (no antibodies prior to vaccination) (n=2,331) | | | | | | | | |
| --- | --- | --- | --- | --- | --- | --- | --- | --- | --- |
|  | Tested 3-5 months ago (n=508) | | | Tested 6-11 months ago (n=1,253) | | | Tested ≥12 months ago (n=570) | | |
|  | n | u% | w% | n | u% | w% | n | u% | w% |
| Definition 1. (≥1 of all symptoms) | 154 | 30.3 | 29.7 | 390 | 31.1 | 28.5 | 183 | 32.1 | 32.3 |
| Definition 2. (different symptoms positives-negatives) | 115 | 22.6 | 20.3 | 280 | 22.3 | 19.8 | 140 | 24.6 | 24.8 |
| Definition 3. (different symptoms positives-negatives plus severity) | 92 | 18.1 | 15.1 | 204 | 16.3 | 14.4 | 113 | 19.8 | 19.2 |
| Definition 4. (WHO) | 62 | 12.2 | 11.5 | 187 | 14.9 | 13.6 | 96 | 16.8 | 18.5 |
| u%=unweighted percentage, w%=weighted percentage. | | | | | | | | | |
